# Long-read nanopore sequencing reveals novel common genetic structural variants in Prader-Willi syndrome and associated psychosis

**DOI:** 10.1101/2022.07.18.22277235

**Authors:** Maximilian Deest, Björn Brändl, Christian Rohrandt, Christian Eberlein, Stefan Bleich, Franz-Josef Müller, Helge Frieling

**Affiliations:** Department of Psychiatry, Social Psychiatry and Psychotherapy, Hannover Medical School, Hannover, Germany; Department of Genome Regulation, Max Planck Institute for Molecular Genetics, Berlin, Germany; Department of Psychiatry and Psychotherapy, Christian-Albrechts University, Kiel, Germany; Institute for Communications Technologies and Embedded Systems, Kiel University of Applied Sciences Kiel, Germany

## Abstract

Prader-Willi syndrome (PWS) is associated with severe hyperphagia, a specific behavioral phenotype and a high risk for developing psychotic episodes. Despite intense research, how genes within the PWS locus contribute to the phenotype remains elusive. In this study, we sequenced the whole genomes of 20 individuals with PWS using long-read nanopore sequencing by Oxford Nanopore Technologies (ONT). We demonstrate that ONT sequencing can resolve the PWS locus by determining the genetic subtype of PWS. Furthermore, we identified several novel structural variants (SV, >30bp) common in all PWS individuals. We are the first to show that the opioid system and the nociceptin/orphanin FQ system may be affected in PWS due to SVs in OPRM1 and OPRL1. Furthermore, we demonstrate that individuals with PWS, especially those with psychosis, exhibit a high burden of SVs in loci with known associations with bipolar disorder, schizophrenia and autism spectrum disorder. Our results challenge the current hypothesis that the PWS phenotype can be mainly explained by the loss of paternally expressed genes on chr15q11.2-13.

## Introduction

Genomic imprinting describes the expression of genes in a parent-of-origin manner. The chromosomal region 15q11.2-13 is subject to imprinting, meaning only the gene copies on the paternal allele are expressed and epigenetically silenced on the maternal allele. Loss of the paternally expressed genes in this region causes Prader-Willi syndrome (PWS) and in around 60% of cases occurs due to a large deletion of approximately 6mb length (delPWS), in around 35% of cases due to maternal uniparental disomy (mUPD), and in up to 5% of cases due to a defect of the imprinting center (ID)^1,2^. Newborns with PWS typically present with hypophagia caused by poor suck leading to a failure to thrive. In later infancy, the hypophagia diminishes and is replaced by pronounced hyperphagia leading to life-threatening obesity if food intake is not controlled. Other hallmarks of PWS are hypothalamic dysfunction, hypogonadism, growth hormone deficiency, an overall delay in language and motor skills and, in almost all cases, a mild to moderate intellectual disability^3^. Certain behavioral traits, such as stubbornness, mood instability, compulsions, skin-picking, and temper outbursts are common in PWS^4^. Six proteins are encoded in the PWS region (*MKRN3, MAGEL2, NDN, NPAP1*, and *SNURF-SNRPN*) and a cluster of several paternally expressed snoRNA genes^5^. To which extent these genes contribute to the phenotype of PWS is not entirely conclusive.

Another hallmark of PWS is the susceptibility to certain mental disorders, especially affective psychosis, autism spectrum disorder, depression, and anxiety with a significantly higher frequency in PWS than in the normal population and even higher than in other causes of intellectual disability^6,7^. There seems to be a genotype-dependent pattern of distribution with individuals with the mUPD and IC subtype showing higher rates of affective psychosis and ASD than individuals with delPWS. in fact, up to 70% of individuals with mUPD PWS experience a psychotic episode once in their lifetime. As the mechanism leading to PWS is different in all subtypes but the result on a genetic level is identical, namely loss of the paternally expressed genes, the mechanisms leading to effective psychosis in PWS remain unclear up to day Hypothesis on a gene dosage effect of UBE3A, which is paternally imprinted and maternally expressed, exist but do not explain why only a fraction of individuals with mUPD exhibit psychosis experience^8^. A possible contributing factor could be genetic variation outside the PWS locus. Long-read sequencing (LRS) such as Oxford Nanopore Technologies (ONT) allows the sequencing of DNA strands of several kilobases in length. Compared to short-read sequencing (SRS), with a read length of 100-200bp, widely used in GWAS studies, LRS detects structural variants (SVs) greater than 30bp more easily and with higher accuracy^9^.

## Results

We sequenced the whole genomes of 20 individuals with a genetically confirmed diagnosis of PWS with the Oxford Nanopore Technologies (ONT) long-read PromethION sequencing platform. The median age of study participants was 26.5 years (range: 12 - 55y, 9 females, 11 males). In 8 cases, PWS was caused by delPWS, in 7 by a mUPD, in 3 PWS was due to an ID and in 2 cases differentiation between UPD and ID did not take place at the time of diagnosis (Suppl. Table 1). In the median, we received 7.39×10^6^ reads per sample with a median read length of 7369.5 bases per read (see Suppl. Table2). The reads were mapped against GRCh38.p13 (hg38) using minimap2^10^, resulting in a median coverage of 29.97 (Suppl. Fig.1).

### Nanopore sequencing can confirm the diagnosis of PWS

First we wondered if we could retrace the genetic PWS diagnostic and determine the genetic subtype by just using ONT sequencing without using other sequencing methods such as MS-MLPA or high-resolution SNP microarrays, which is the common approach for genetic testing. In all cases of PWS, due to the loss of the methylation pattern on the paternal allele, methylation frequency of CpGs is higher at the PWS locus than normal. We called methylation frequencies of CpGs over the whole chr15 including the PWS locus using nanopolish^11^ and compared methylation frequencies with a control sample (HG01109 from the T2T diversity panel). All PWS individuals show a higher mean methylation rate (mean 71.78%, SD: 10.1%) across the PWS locus than the control sample (mean 44.91%, SD: 21.5%), in line with what was expected. (Fig 1. A) (Methylation data is found in Suppl. Table 3). Several genes (e.g. *MKRN3, MAGEL2, NDN, PWRN1, SNURF-SNRPN, UBE3A*) within the PWS locus underlie genetic imprinting. Thus, the expression of those genes is epigenetically silenced on one parental gene by methylation. At the PWS locus, *MKRN3, MAGEL2, NDN, PWRN1, SNURF-SNRPN* are highly methylated on the maternal gene and just expressed on the paternal allele, whereas it is the other way around with *UBE3A*. We analyzed allele-specific methylation by calling single nucleotide variants (SNV) at chr15 with longshot^12^. The sets of SNPs for each individual were used to phase the alleles by haplotypes using whatsHap^13^. Methylation data were then separated for each haplotype block. As expected in PWS, methylation rates were similarly high at both alleles at genes that are imprinted compared to two differentially methylated alleles in the control sample (Fig. 1B).

**Figure 1.**
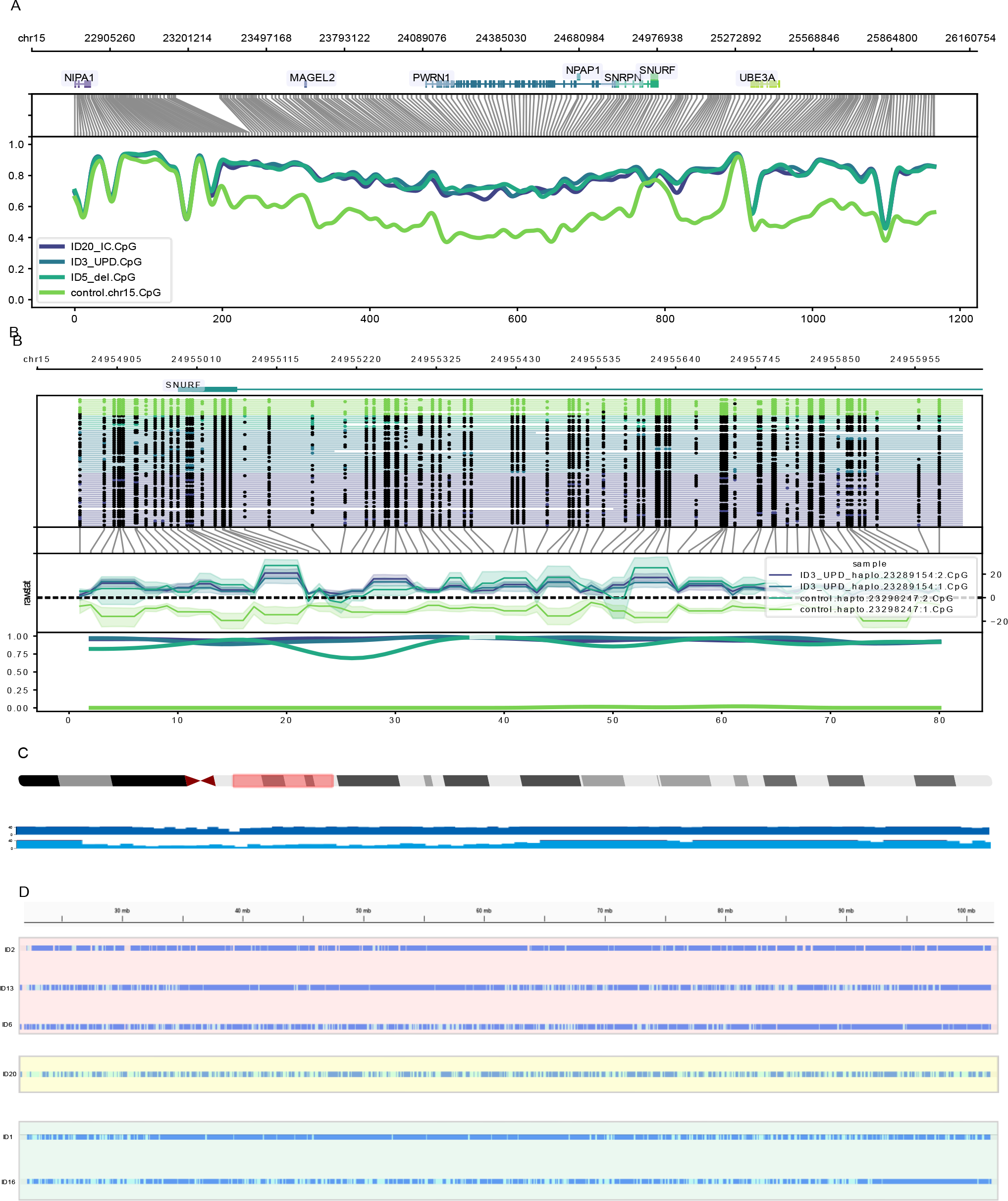
A) Methylation plotted over the PWS region with light green being the control sample, purple imprinting center defect, dark blue UPD, and light blue delPWS. The PWS samples show significantly higher methylation across the imprinted genes due to the absence of the paternal allele in PWS. Methylation of control samples and PWS samples converge at the point of UBE3A, a gene that is highly methylated at the paternal allele but not the maternal one. B) Methylation plotted over the imprinted gene SNURF. In PWS both alleles are highly methylated due to the loss of the paternal allele, whereas in the control samples, the alleles are differentially methylated. C) Coverage plot against hg38. Showing a large deletion of chr15q11-q13 in a PWS individual with type II deletion (ID5) in light blue. Darkblue is ID20 (IC) as a control. D) Fig: Visualization of the genotypes over chr15 called by longshot.Size of the region in mb as a comparison. Regions with LOH > 8Mb are indicators of isodisomy. Dark blue colors refer to homozygosity (GT 1/1) and light blue to heterozygosity (GT0/1, 1/0). Red panel: If both chromosome 15s are inherited from the same chromosome 15 from the mother (isodisomy) we expect homozygosity over the whole chromosome (ID2). If only parts of the chromosome are identical, we expect large regions of homozygosity (ID13 & ID6). Yellow panel: ID20 (IC defect) shows heterodisomy with no regions >8Mb of LOH as a control Lowest panel (green) are the samples ID1 & ID16, with a previously unknown subtype. Large regions of LOH indicate a UPD subtype.

### Nanopore sequencing allows the determination of the genetic subtype in PWS

Proof of higher methylation over the PWS region confirmed the diagnosis of PWS in all samples. In the next step, we determined the genetic subtype. First, we resolved the large deletions of chr15q11.2 - chr15q13 on the paternal allele in the case of delPWS. Using standard variant callers such as cuteSV^14^ was not possible in this case due to two reasons: 1) the deletions in PWS only are present on one allele and 2) the deletions are of several Mbps length and thus exceed the current maximum read length, even of long-read sequencing methods such as ONT. However, as the number of reads covering a region is proportional to the number of DNA strands present in a sample, read coverage at a large deletion is lower than outside the deletion. Read coverage was calculated using deepTools and can be manually inspected using standard genome browsers such as JBrowse^15^. Samples with a delPWS show a large region of lower coverage at the region of deletion (Fig.1C).

In the case of mUPD or ID, no large deletion is detectable. Maternal UPD refers to the mother’s inheritance of both copies of chr15. This can be due to 1) a total isodisomy (two copies of one chr15 from the mother) due to a meiosis II error, 2) segmental isodisomy (only parts of one chr15 from the mother) due to crossover(s) in meiosis I and 3) heterodisomy, (inheritance of both homologous chr15 from the mother) due to the fact that no crossover events occurred. Isodisomy and segmental isodisomy are detectable by large regions of loss of heterozygosity (LOH), which have a minimum length of 8*mb*. We used the SNP data produced by longshot^12^ with a minimum allele frequency of 15 as a cutoff to determine the genotypes. Large regions with a genotype of 1/1 indicate LOH, whereas genotypes 0/1 or 1/0 indicate heterozygosity. This method allows us to detect both forms of isodisomy (total and partial). Interestingly the size and locations of regions of LOH differ between samples (Fig.1D) as reported previously^5^. The location of large regions of LOH can have a significant impact if the mother carries a recessive disease-specific allele in that region.

Heterodisomy can not be detected without parental DNA to check for biparental inheritance of chr15. However, if no large deletion and no large regions of LOH are detected, PWS could be due to errors in genetic imprinting (ID), either caused by a microdeletion in the imprinting center or an epimutation without changes in the DNA sequence. The imprinting center refers to a 4.3 kb region spanning the SNRPN promoter, exon 1, and intron 1. Manually inspecting the alignment files of the three samples with IC revealed a 20kb deletion (ID10, ID20, chr15: 24930934-25132606) and a 29kb deletion (ID18, chr15:24834729-25126076) spanning the imprinting center (Supp. Fig2). Thus, we could also detect IC defects caused by microdeletions.

For two of our samples (ID1 & ID16), we knew that the genetic subtype was not a delPWS but differentiation between UPD or ID did not take place at the time point of genetic counseling. We used our above-described approach and could detect large regions of LOH in both samples, indicating that an isodisomy was the cause of PWS (Fig. 1D).

To distinguish between the last two possibilities causing PWS, namely a maternal uniparental heterodisomy and an epimutation, DNA from both parents is needed to check for biparental inheritance (in the case of an epimutation). About 60% of PWS cases arise through a large deletion, 36% through an mUPD, and 4% through an IC defect. Of the mUPD cases, around 70% are caused by isodisomy and 10% of IC defects are caused by a microdeletion^16^. Thus, ONT sequencing can sufficiently determine the genetic subtype in 85.6% of cases without using parental DNA. In a clinical setting, ruling out or detecting a microdeletion in the IC is of importance, as in the case of a microdeletion the occurrence risk is up to 50%.

### Novel structural variants shared in all PWS individuals

ONT sequencing allows the reliable detection of SVs^9^. We wondered if there are any SVs outside the PWS locus which may contribute to the PWS phenotype. Therefore we used cuteSV^14^ to call SVs >30bp in length. This resulted in a median call of 28,123 SVs per sample, with a median number of 15,166 (53.93%) insertions and 12519 (44.52%) deletions. The rest are duplications, inversions, and translocations. The majority of SVs were located within introns (64.1%) or intergenic locations and only a minority of SVs were located in exons (0.73%), the 3’UTR (0.21%) or 5’UTR-(0.04%). Next, we were interested in which SVs are shared between all samples. As proper variant calling is coverage dependent we decided to only include samples with a median coverage of at least 20 for further analysis. Thus we had to drop ID 2 (median coverage 13.71) and ID 18 (median coverage 18.5). We used JASMINE^17^ to merge SVs present in all samples. Merging of the variants resulted in a total of 4220 SVs present in all samples (1325 DEL, 2857 INS, and 38 others). For pairwise comparison see Fig2. (pairwise correlation matrix: Suppl. Table04).

**Figure 2.**
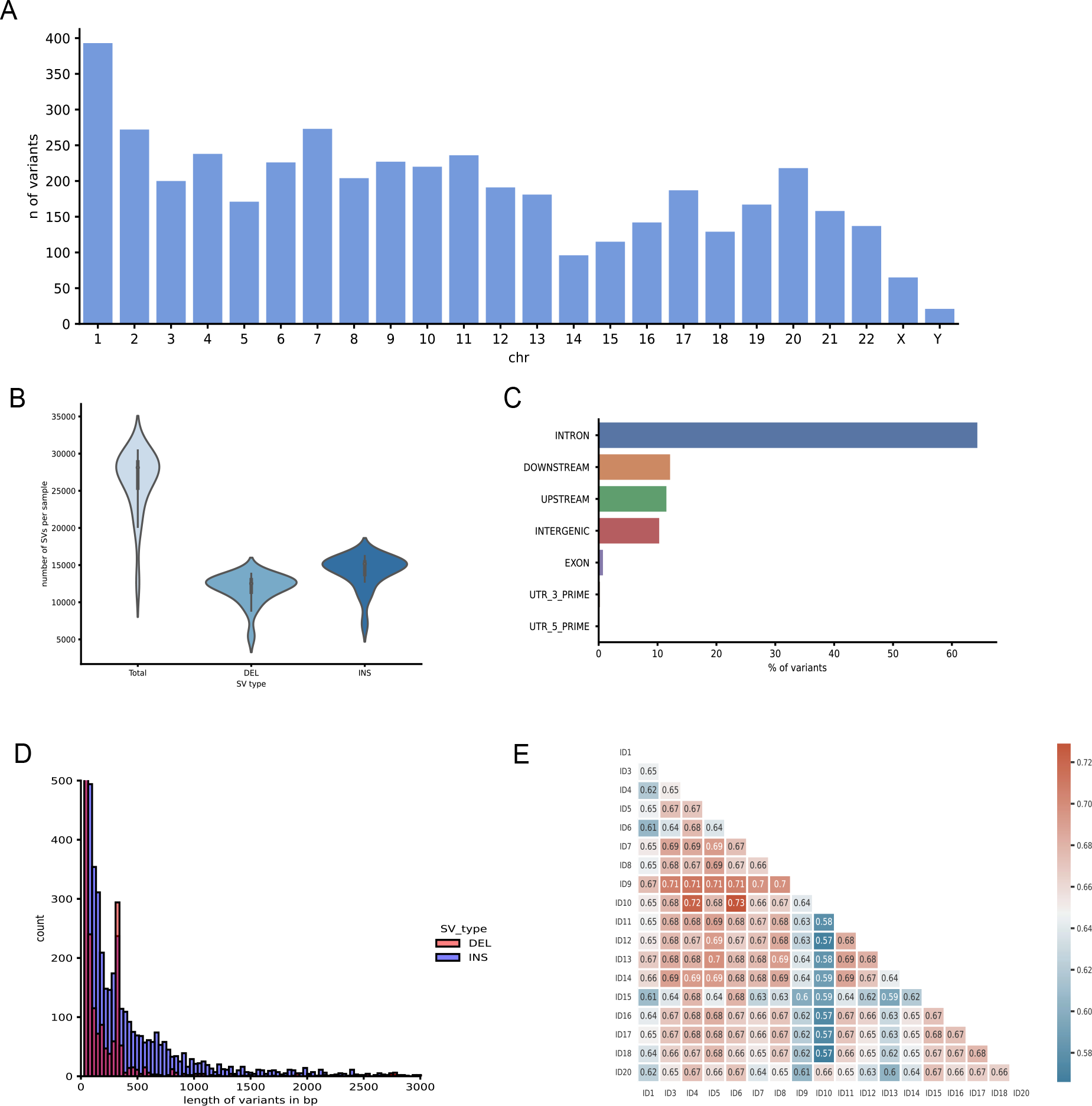
Characteristics of SVs. A) shows a barplot of variants detected per chromosome. B) violin plot of type of variants found C) Barplot of numbers of variants located within which parts of the genome (e.g. exon, intron, intergenic) D) Histogram of length distribution of SV types deletion and insertions E) Heatmap of pairwise-correlation of SVs found within each sample.

**Figure 3:**
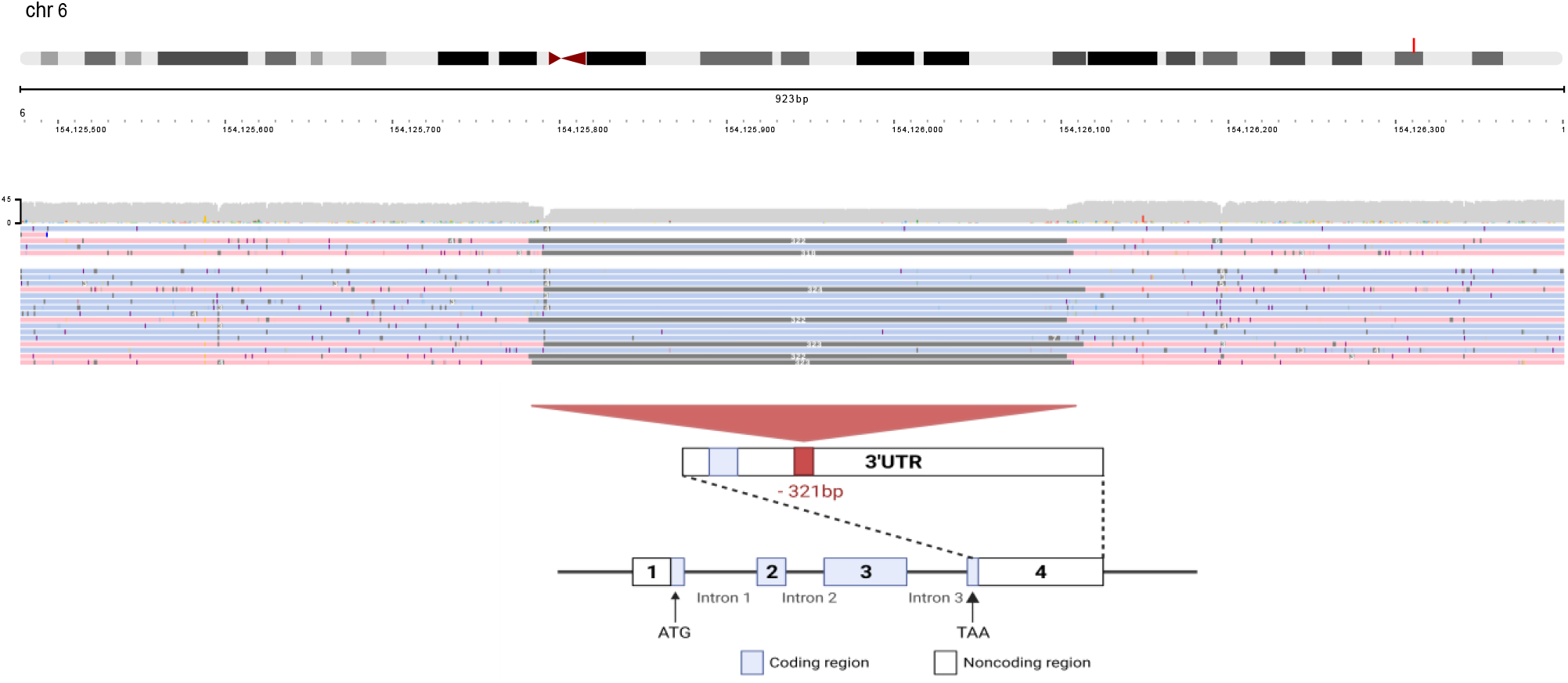
Visualization of the -321bp deletion within the 3’UTR of OPRM1. OPRM1 is located at chr6:154,010,496-154,246,867. At the top is the ideogram of chr6 with the respective localization of OPRM. In the middle are aligned reads colored by haplotypes indicating the alleles. As OPRM is monoallelic, the deletion only spans one allele. At the bottom is a schematic simplified figure of the OPRM gene structure and the localization of the deletion within the gene

**Figure 4:**
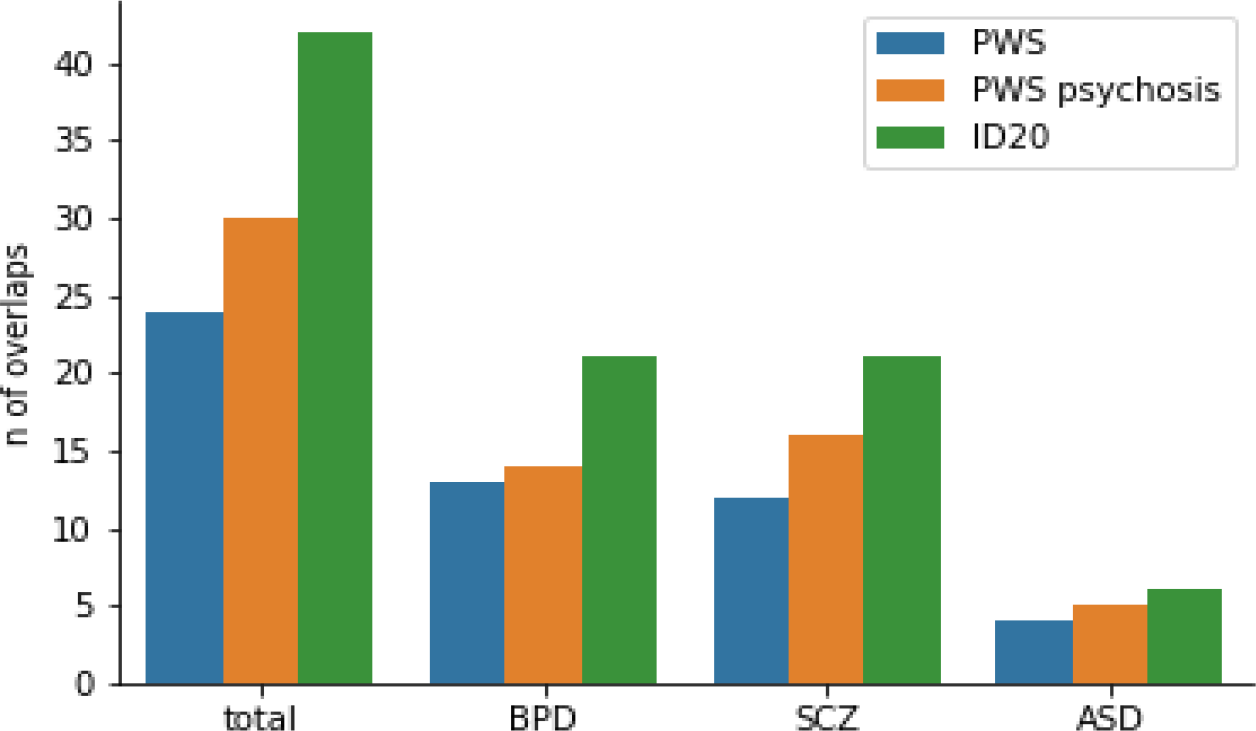
Genes impacted by structural variants overlapping with genes found in GWAS studies for BPD, SCZ, and ASD.

SVs were annotated using AnnotSV^18^ and manually checked for impact and relevance. We were highly interested in SVs within genes or regions which might contribute to certain domains of PWS, like intellectual disability or the high prevalence of psychotic symptoms or autism. Interestingly we found several SVs in genes located within chr22q11.2. Genetic variation in this region leads to other neurodevelopmental disorders: The chr22q11.2 duplication syndrome is associated with an intellectual or learning disability, developmental delay, slow growth leading to short stature, and a weak muscle tone, and neuropsychiatric comorbidities such as observed in PWS^19^. A deletion in this region, spanning around 3 Mb leads to the DiGeorge syndrome, associated with the development of psychotic symptoms and ASD. In our cohort, we found a -65 bp deletion within the 5’UTR of COMT (Catechol-O-methyltransferase). COMT is one of the major enzymes involved in the metabolic degradation of catecholamines. Studies showed that COMT possibly contributes to obsessive-compulsive disorder (OCD)^20^ and schizophrenia^21,22^. Individuals with PWS often exhibit repetitive and ritualistic behavior as seen in OCD. Interestingly it could be recently shown that a deletion within the 5’UTR of COMT alters the expression levels^23^. Furthermore, we found two insertions (202bp in exon 1, 1320bp in intron1) in RTN4R (Reticulon 4 receptor). It plays a role in regulating axon regeneration and neuronal plasticity in the adult central nervous system and variants within RTN4R are associated with schizophrenia^24^. Another deletion in this region was found in intron 7 of LZRT1 (leucine zipper like transcription regulator 1), a gene that can be truncated in Noonan-Syndrome^25^. Interestingly, Noonan-Syndrome is also associated with short stature and, as PWS, with hypophagia in the first two years of life with poor suck and a failure to thrive. Another interesting SV is a -37bp deletion in 3-UTR of HHIP (Hedgehog-interacting protein), a gene associated with body height^26^.

### OPRM1 3’UTR is truncated in Prader-Willi syndrome

We found a -321bp deletion in the OPRM1 (opioid receptor mu 1) gene. OPRM1 encodes the mu opioid receptor (MOR), one of at least three opioid receptors in humans^27^. The deletion is located within the 3’UTR of exon4 of OPRM1 (chr6: 154,125,782-154,126,105). OPRM1 is a monoallelic gene undergoing heavy alternative splicing. Only having one copy of OPRM1 is unusual for a receptor with this many ligands and functions. The hypothesis is that the single copy of the OPRM1 gene creates multiple mu-opioid receptor splice variants or isoforms through alternative pre-mRNA splicing^28^. The 3-UTR of OPRM1 is known to have extensive regulatory functions by binding multiple transcription factors^29^. Since 3’UTRs affect gene expression, a large deletion within this region possibly leads to dysregulation of the expression of OPRM1. The mu opioid receptor (MOR) is the principal target of endogenous opioids like beta-endorphine or enkephaline as well as exogenous opioids. It plays an important role in the development of substance dependences (e.g. nicotine, opioids, and alcohol) as it modulates dopaminergic neurotransmission within the reward system. MOR functioning has been implicated with a wide variety of behaviors, ranging from emotional eating^30^ over impulsivity^31^ to social behavior^32^.

Moreover, we found a -267bp deletion in the 5-UTR region of intron 1 of OPRL1 (opioid related nociceptin receptor 1). The protein encoded by OPRL1 functions as a receptor for the endogenous, opioid-related neuropeptide, nociceptin/orphanin FQ. This peptide is involved in the modulation of stress, addiction, mood, anxiety, obesity, and binge-eating behavior^33^.

### Individuals with PWS share genetic variants within gene loci highly associated with bipolar disorder, schizophrenia, and autism spectrum disorder

PWS is associated with a high risk for the occurence of psychotic symptoms and autism spectrum disorder. Symptoms of psychotic disorders overlap those observed in bipolar disorders as well as schizophrenia and often are entitled affective psychosis. Multiple genome-wide association studies on BPD, SCZ and ASD have been performed in the past, identifying several genes possibly contributing to these disorders. We wondered if a fraction of these genes are also affected in PWS and individuals with PWS and psychosis. In our cohort, 4 individuals had at least one confirmed episode of psychotic illness. We merged all SVs across those 4 individuals resulting in a total of 10,558 SVs shared between them. To reduce the number of potential variants for further analysis we used the following approach. If we would have selected only those SVs which are not present in the merged SV set of all individuals, we would probably lose promising SVs as maybe some of the other individuals also share some of these variants, but have not yet developed a psychosis. As one of the individuals with psychosis also has a sibling within the study population who does not have psychosis, we used the siblings set of SVs as a “control group’. As the sibling was already over 50 years old at the time point of blood sampling and never had a psychotic episode in it’s life, it is highly unlikely that he/she will develop a psychosis in the future. The psychotic group shared 8773 SVs with the sibling, making those variants very unlikely to contribute to the development of psychotic illness. However, 1785 SVs were only present in the psychotic group but not in the sibling, thus we decided to further investigate them.

We wondered if impacted genes in our cohort overlap with loci associated with mental disorders found in recent GWAS studies. Therefore we compared the most significant results of recent GWAS studies investigating BPD^34^, SCZ^35,36^, and ASD^37^. This resulted in a total set of 155 genes (62 BPD, 70 SCZ, 27 ASD, 4 of them overlapping disorders). A total number of 24 out of these 155 genes (13 BPD, 12 SCZ, 4 ASD) were impacted in any way (deletions/insertions, in introns or exons) in all PWS samples. In the group of PWS with psychosis 30 genes (14 BPD, SCZ 16, ASD 5) overlapped. Last we looked at one individual with psychosis alone, resulting in 42 overlapping genes (21 BPD, 21 SCZ, 6 ASD) (Find all affected genes in Suppl. Table05). This indicates that the burden of risk-associated loci is generally high in PWS but even higher in PWS with psychosis. Looking at an individual level, the burden increases even more. Thus we conclude that the risk of developing psychosis in PWS is not associated with a single gene alone but by the combination of several affected genes. The six genes present in all individuals with PWS and psychosis but not in the merged file with all SVs shared in PWS are listed in Table2. We found two deletions within intron 1 of GRIK3 (Glutamate receptor ionotropic, kainate 3). GRIK3 encodes a glutamate receptor binding N-methyl-D-aspartate and it has previously been described that SNVs located in intron 1 of GRIK3 are associated with schizophrenia^38^

**Table1:** SVs within genes which might contribute to the phenotype of PWS found in all samples

| Gene | SV type | SV length | Location | chr: start - end |
| --- | --- | --- | --- | --- |
| COMT | DEL | -65 | intron 1 | 22:19959795 - 19959865 |
| RTN4R | INS | 200 | exon 1 | 22: 20268314 - 20268316 |
| RTN4R | INS | 1320 | intron 1 | 22: 20251916 - 20251934 |
| LZTR1 | DEL | -36 | intron 7 | 22: 20989699 - 20989739 |
| HHIP | DEL | -37 | exon 13 | 4: 144742961 - 144743000 |
| OPRM1 | DEL | -321 | exon 4-3'UTR | 6: 154125782 - 154126105 |
| OPRL1 | DEL | -267 | intron 1 | 20: 64090771 - 64091069 |

**Table2:** SVs in Genes associated with SCZ, BPD, ASD shared by all PWS individuals with psychosis but not present in the overall population

| Gene | SV type | SV length | Location | chr: start - end |
| --- | --- | --- | --- | --- |
| ANK3 | INS | 137 | intron 8 | 10: 60226231 - 60226506 |
| GOLGA6L4 | DEL | -31 | intron 9 | 15:84509704 - 84509749 |
| GRIK3 | DEL | -284 | intron 1 | 1: 36958130 - 36958428 |
| GRIK3 | DEL | -46 | intron 1 | 1: 36959923 - 36959973 |
| MROH5 | INS | 35 | intron 6 | 8: 141488217 - 141488226 |
| NGEF | INS | 120 | intron 1 | 2: 232993170 - 232993188 |
| TSNARE1 | INS | 359 | intron 12 | 8: 142260999 - 142261007 |

## Methods & Materials

### Human subjects

This study adhered to the Declaration of Helsinki and was approved by the local Ethics Committee of Hannover Medical School (Nr. 8129_BO_S_2020), Hannover, Germany. All participants and/or their legal representatives gave their written informed consent for participation after the nature of the procedures had been fully explained. This study included 20 individuals with a genetically confirmed diagnosis of Prader-Willi syndrome. Participants were recruited at the Outpatient Department for Mental Health in Rare Genetic Disorders, part of the Department of Psychiatry, Social Psychiatry and Psychotherapy of Hannover Medical School, Hannover, Germany. Recruitment took place between 2019 and 2021. Only individuals with a genetically confirmed diagnosis of PWS were included, Genetic diagnosis was performed at licensed genetic laboratories and written results were present. However, genetic subtyping was not mandatory and did not take place in two participants. At baseline, a comprehensive interview with the participant and their caregivers (e.g. parents) did take place in order to gather data on demographics and medical history. Assessment of psychotic episodes did take place via checking the medical history when the episode was in the past, and by checking the current symptoms at the time point of the study by using the ICD-10 criteria.

### Blood collection & DNA extraction

EDTA blood was collected in all participants. DNA was extracted from blood directly after sampling by the Hannover Unified Biobank using the Hamilton ChemagicStar (Hamilton Germany Robotics, Graefelfing, Germany) and the chemagicStar DNA-Blood1k kit (PerkinElmer chemagen Technology, Baesweiler, Germany).

### Sequencing

DNA was then used for the Genomic DNA by Ligation (SQK-LSK109) library preparation (ONT, UK) according to the published protocol with minor adjustments: Up to 63μg of DNA was used for each library preparation; DNA Repair and End-prep reaction incubation times were increased to 20 minutes at 20 °C and 10 minutes at 65 °C; End-prep reaction AMPure XP bead cleanup elution step was performed for 25 minutes at 60 °C with flicking every five minutes; Adaptor ligation was performed for 30 minutes at room temperature; AMPure XP beads were then incubated for 30 minutes at 48 °C with flicking and washed with “Long Fragment Buffer;” 80% EtOH was used throughout for all washing steps. Sequencing was performed on a PromethION instrument using R9.4.1 flow cells. Washes were performed after 20-24 hours using the Flow Cell Wash Kit (EXP-WSH003) and reloaded with a fresh library for a total run time of 72 hours.

### Long-read whole genome sequencing analysis

Raw nanopore reads (fast5 format) were basecalled using Guppy Basecalling Software v6.1.2 (Oxford Nanopore Technologies plc.) using the default settings. Reads were aligned to the Genome Reference Consortium Human Build 38 patch release 13 (GRCh38.p13, hg38) using minimap v2.2^10^ with the flags *-L, –MD, -ax map-ont*. Coverage statistics were calculated using mosdepth^39^.

Methylation calls were determined using nanopolish ^11^v0.13.2 with the recommended settings. As a control sample we used HG01109, a male individual sequenced by the T2T Diversity Panel (available at: https://github.com/human-pangenomics/hpgp-data). Methylation frequencies were calculated for the PWS region (chr15:2356500 - 2533500) in 1000bp bins. Methylation frequencies for all samples over the PWS locus can be found in Suppl. Table3. Methylation plot was generated using the *–region* function of methylartist^40^ v1.0.5. To phase the methylation data for each allele we used the following approach. First, single nucleotide variants were called at chr15 using longshot^12^ v0.4.1 with a *–min_alt_count* of 15. To filter out sides with genomic features such as copy number variations, which can be problematic for proper variant calling, we used the *-A* flag. With the *-A* flag longshot estimates the mean read coverage and sets the max coverage to *mean_coverage +5*sqrt(mean_coverage)*. The outputted Variant Call file (VCF) file of longshot was used as an input for whatsHap^13^ v0.18 to phase the BAM files containing the aligned reads. The default settings of whatsHap were used. The phased methylation data were then plotted with the locus function of methylartist with the *–phased* flag.

For identification of the larger deletions inside the PWS locus we generated a coverage track using the bamCoverage function of deepTools^41^ v.3.5.0. The coverage was calculated as the number of reads per bin, with a default bin size of 50. Visualization of the coverage tracks was done using JBrowse2^15^.

To assess the deletions spanning the imprinting center in ID we manually inspected the BAM files produced by minimap2 using JBrowse2. The PWS SRO (smallest region of overlap) consists of SNRPN (small nuclear ribonucleoprotein polypeptide N) promoter, exon 1 and intron 1 (located at chr15:24,871,386 - 24,874,124 as referred to the USCS genome browser). Deletions could be detected by an abrupt drop of coverage and soft-clipped reads (Supp.Fig2).

Determining of isodisomy, heterodisomy, or partial isodisomy in the UPD cases was performed as follows. Longshot was used as described above to determine SNVs over the whole chr15 for each sample separately. The resulting VCF files were checked for the genotypes (GT). A GT of 0|1 or 1|1 indicates heterozygosity, a GT of 1|1 indicates homozygosity. SNV in dense clusters of potential false calls, e.g. due copy numbers or mapping issues were filtered out. VCF files were then loaded into the IGV genome browser and the track was colored by GT. Tracks were manually inspected to determine isodisomy (nealy full GT 1|1), heterodisomy (GT 0|1 or 0|1 over the whole chromosome). Regions of LOH had to be >8mb for isodisomy.

#### Variant calling

Calling of structural variants (>30 bp) was done using cuteSV14 vXY with default settings resulting in one VCF per sample. SVs across samples were merged using JASMINE17 v1.1.5 resulting in one VCF file with SVs present in all samples. JASMINE uses a minimum spanning forest algorithm and takes SV type (such as deletion, insertion, inversion, translocations), position information (chr, start, end) and strand information into account. SVs were annotated using annotSV18. SVs were manually validated for their impact by using public available databases such as OMIM or pubmed.

For comparison of genes affected by any kind of structural variation in our cohort against identified high-risk loci for bipolar disorder (BPD), schizophrenia (SCZ), and autism spectrum disorder (ASD) we used the following sets: For BPD we used 62 genes from Mullins et. al.34, for SCZ we used genes found in Kirov et. al.36 and overlapping with genes from Lee et. al.35, resulting in 70 genes and for ASD we used 27 genes from Giovani et. al.37, resulting in a total of 155 genes (4 genes overlapped between disorders).

## Data Availability

All data produced in the present study are available upon reasonable request to the authors

